# Shortened blastocyst vitrification achieves live birth rates comparable to standard protocols: an analysis of 3168 cryotransfers

**DOI:** 10.64898/2026.06.11.26354186

**Authors:** E.R. Ebinger, H. Misurac, C. Lynch, V.B. Desai, N. De Rosa, R. Vassena, P. J. Atkinson

**Affiliations:** Adora Fertility, Level 2 Westside Private Hospital, 30 Morrow Street, Taringa QLD 4068 Australia; Adora Fertility, 28 Foveaux Street, Surry Hills, NSW 2010 Australia; CooperSurgical Fertility Solutions, Trumbull, CT, USA

**Keywords:** vitrification, fast protocol, ICSI, IVF, blastocysts

## Abstract

**Study question:** Do shortened blastocyst vitrification and warming protocols provide comparable live birth rates (LBR) and obstetrical and perinatal outcomes to traditional vitrification and warming protocols?

**Summary answer:** Shortened vitrification and warming protocols provide comparable LBR, obstetric and perinatal outcomes to traditional protocols.

**What is known already:** Embryo viability following cryopreservation is dependent on blastomere survival and functional integrity, both impacted by ice crystal formation and osmotic gradients. Recent innovations in cryopreservation challenge the need for stepwise dehydration and rehydration protocols. While one-step “fast” blastocyst warming protocols seem to provide equivalent clinical outcomes to traditional “slow” protocols, fewer studies investigate whether blastocyst dehydration rates can be similarly increased. A thorough safety and effectiveness evaluation remains necessary for both treatment success and offspring health.

**Study design, size, duration:** Retrospective, multicentric, consecutive cohort analysis. Three groups were established for analysis from survival to live birth – traditional vitrification/multi-step warming (S/S, n=751 blastocysts warmed) short vitrification/multi-step warming (F/S, n=991), and short vitrification/one-step warming (F/F, n=1593). S/S was set as the control group for statistical purposes. We modelled the relationship between “fast” versus “slow” protocols and outcomes with Generalized Additive Models, and linear and logistic regressions where appropriate. Two-tailed chi-square with Yates correction was used to examine pregnancy loss and obstetrical and perinatal outcomes; p<0.05 was considered significant.

**Participants/materials, setting, methods:** Three clinics within a network participated in this retrospective consecutive cohort study, with cycle data collected for 3603 warmed blastocysts resulting in 3168 frozen blastocyst transfers in 2170 patients between 2023 and 2025. All cycles were with the patients’ own gametes and without preimplantation genetic testing. All blastocysts were collapsed before vitrification, which was either a traditional “slow” protocol (Sydney IVF, Cook Medical) or an amended 2.5-minute “fast” protocol (SAGE^TM^, CooperSurgical). Warming was either a traditional “slow” multi-step protocol or a “fast” one-step 1minute protocol in 1.0M sucrose (both SAGE^TM^, CooperSurgical).

**Main results and the role of chance:** The overall LBR was 33.1% (1050/3168) with female age 35.2±4.5 (range:22-46). Overall, all reproductive outcomes, including live birth rates, as well as obstetric and perinatal outcomes, were comparable across all groups (all p>0.05). This analysis included a total of 3603 blastocysts warmed, and 3335 blastocysts transferred in 3168 procedures for 2170 unique patients. A total of 751 blastocysts were transferred to 555 patients in group S/S, 991 were transferred to 734 patients in the group F/S, and 1593 were transferred to 1126 patients in group F/F. Blastocyst survival rates were 99.1%, 99.0% and 99.3% for groups S/S, F/S and F/F respectively. The biochemical pregnancy rates were 46.2% for S/S, 49.6% for F/S, and 43.9% for F/F. Clinical pregnancy rates were 43.1% for group S/S, 46.1% for group F/S, and 40.8% for group F/F. Live birth rates were 33.6% for group S/S, 36.8% for group F/S, and 30.6% for group F/F.

Interestingly, women 35yrs or older at vitrification (n=1715 transfers), and transfers with lower quality embryos seemed to profit from a F/S strategy.

**Limitations, reasons for caution:** Factors affecting the results may be unaccounted for by the study retrospective nature. Further, all clinics included in the study operated under the same clinical protocols, which may limit the generalizability of results.

**Wider implication of the findings:** Overall, shortened, “faster” vitrification and warming protocols provide comparable reproductive outcomes to traditional ones. Longer term marker of embryo health such as perinatal outcomes are also comparable, indicating overall safety of these shortened procedures.

## INTRODUCTION

The development of blastocyst vitrification has been critical in establishing highly effective cryopreservation programs, improving the cumulative live birth rate of treatments and allowing for the postponement of embryo transfers with minimal risk of embryo loss during warming. Global data indicate that frozen embryos transfers (FET) account for 61.8% of autologous transfers without preimplantation genetic testing (PGT). At the same time, “freeze-all” cycles without fresh transfer have increased from 24.6% to 38.4% (Dyer *et al*., 2025).

Recent reassessment of longstanding embryo cryopreservation protocols has established the non-inferiority of a one-step 1 minute warming protocol in 1.0M sucrose with no observable reduction in embryo viability (Manns *et al*., 2022; Liebermann *et al*., 2024; Karagianni *et al*., 2025; Ebinger *et al*., 2026).

Equilibration, on the other hand, is the initial and longest step in traditional vitrification. During equilibration, the embryo shrinks as it dehydrates, then swells again as permeating CPAs replace lost water and an osmotic equilibrium is attained. Next, embryos are exposed for 1 minute to a solution with a higher concentration of permeating CPAs, allowing for further dehydration and CPA permeation. Data on shortening the equilibration step to as low as 2 minutes are promising (Kaskar *et al*., 2023; Arkfeld *et al*., 2025) but still limited. Overall, these shorter protocols challenge the need for traditional extended dehydration and rehydration stages, as well as the time required for the exchange of water and permeating CPAs. Specifically, reducing the equilibration time during vitrification and eliminating the stepwise introduction of non-permeating CPAs during warming increases the rate of both dehydration and rehydration.

Importantly, thermal dynamics remain unchanged during shorter cooling and warming protocols. The cooling rate, and more so the warming rate, are of vital importance to maintain blastocyst viability. The relationship between thermal dynamics and membrane permeability means that an inadequate cooling rate can result in suboptimal CPA penetration (Fahy and Rall, 2007; Seki and Mazur, 2008). Insufficient cooling and warming rates can also cause inter- and intracellular ice crystal formation and damage to membranes and intracellular structures, leading to reduced survival and viability (Muldrew and McGann, 1994). Maintaining cooling and warming rates is therefore key to the success of shortened protocols.

Here, we set out to compare the efficacy and safety of three combinations of shortened (“fast”) and traditional embryo vitrification and warming protocols up to live birth and including obstetric and perinatal outcomes. Additionally, we examine embryo cohorts normally associated with poorer cycle outcomes, including lower embryo quality and developmental rate, presence of male factor infertility, and advanced maternal age.

## MATERIALS AND METHODS

### Study design and population

Retrospective consecutive cohort study at 3 clinics in the Adora Fertility network from September 2023 to April 2025. In May 2023 the laboratories included in the study switched from traditional vitrification to a shortened vitrification protocol of 2 minutes in equilibration solution and 20-30 seconds in vitrification solution. This was followed by a change in warming protocol in July 2024 from a traditional, multi-step protocol to a 1-minute one-step protocol. All cycles included in the analysis were performed with the patients’ own gametes, and without preimplantation genetic testing. Demographic and cycle characteristics are detailed in **Table 1**. The study received ethical clearance from GSAC (approval code:EMB-ADO-181225).

**Table 1:** Demographic and cycle characteristics in the reference group (S/S = traditional vitrification/multi-step warming) and control groups (F/S = short vitrification/multi-step warming, F/F = short vitrification/one-step warming). Numerical results reported as mean ± standard deviation (range). Categorical variables presented as n (%). BMI = Body Mass Index. ICSI = Intra Cytoplasmic Sperm Injection. FET = Frozen Embryo Transfer.

|  | S/S | F/S | F/F |
| --- | --- | --- | --- |
| Patients (n) | 570 | 756 | 1218 |
| Female age (years) | 34.31 ± 4.62<br>(22.25–45.38) | 35.27 ± 4.27<br>(22.33–44.13) | 35.40 ± 4.49<br>(22.05–45.99) |
| Female BMI (kg/m<sup>2</sup>) | 25.04 ± 4.48<br>(15.81–40.35) | 25.10 ± 4.43<br>(16.71–39.76) | 25.08 ± 4.58<br>(16.53–39.76) |
| Male age (years) | 36.61 ± 6.39<br>(21.83–64.22) | 37.49 ± 5.98<br>(22.83–68.38) | 37.58 ± 5.98<br>(22.08–70.97) |
| Sperm motility (a+b;%) | 32.25 ± 27.82 | 29.51 ± 24.86 | 27.10 ± 24.72 |
| Sperm concentration (mill/mL) | 46.68 ± 41.73 | 41.59 ± 38.91 | 39.94 ± 37.20 |
| ICSI (% of cases) | 37.96% | 42.87% | 42.73% |
| Total embryos warmed | 775 | 1053 | 1775 |
| Total embryos transferred | 751 | 991 | 1593 |
| Top quality embryos (group 1) | 29.69% | 34.01% | 28.69% |
| Total transfer procedures (n) | 714 | 940 | 1514 |
| SET | 94.82% | 94.57% | 94.78% |
| Day 5/6 | 623/91 | 864/76 | 1362/152 |

### Ovarian stimulation and oocyte retrieval

Patients underwent controlled ovarian hyperstimulation (COH) according to the clinic protocols and established good clinical practice. Day 1 blood tests were required, demonstrating E2<200, P4<3 and FSH<15 to start stimulation. Clinician preference and clinical indication determined the stimulation protocol and gonadotrophin used. The antagonist protocol was used for most cycles, with agonist and flare protocols employed where clinically indicated. A range of gonadotrophins were used, including rFSH-alfa (Gonal-f^®^, Merck Healthcare KGaA, Darmstadt, Germany and Bemfola^®^, Gedeon Richter Plc., Budapest, Hungary), rFSH-beta (Puregon^®^, N.V. Organon, Oss, The

Netherlands), rFSH-delta (Rekovelle^®^, Ferring Pharmaceuticals, Saint Prex, Switzerland), combined rFSH-alfa and rLH-alfa (Pergoveris^®^, Merck Healthcare KGaA, Darmstadt, Germany), and HP-hMG containing FSH and hCG (Menopur^®^, Ferring Pharmaceuticals, Saint Prex, Switzerland). Ganirelix (Orgalutran^®^, N.V. Organon, Oss, The Netherlands) and cetrorelix (Cetrotide^®^ Merck Healthcare KGaA, Darmstadt, Germany) were used in agonist cycles and triptorelin (Depcapeptyl^®^, Ipsen Ltd., London, UK) or r-hCG (Ovidrel^®^, Merck Healthcare KGaA, Darmstadt, Germany) as the ovulation trigger. Where clinically indicated, r-hLH-alfa (Luveris^®^, Merck Healthcare KGaA, Darmstadt, Germany) was added to the stimulation protocol.

Ovulation was triggered when two or more follicles ≥18mm were detected. Transvaginal oocyte retrieval was performed under heavy conscious sedation using a single lumen needle (Wallace^®^, CooperSurgical, Trumbull, CT, USA or COOK^®^, Cook Medical Australia, Brisbane, QLD, Australia). Cumulus oocyte complexes (COCs) were collected in Quinn’s Advantage™ Protein Plus Fertilization (HTF) Medium (CooperSurgical, Trumbull, CT, USA) and then washed in gassed Origio Sequential Fert^TM^ medium (CooperSurgical, Trumbull, CT, USA). They were finally then placed into Nunc^TM^ 4 well culture dishes (Thermo Fisher Scientific, Scoresby, VIC, Australia) in Origio Sequential Fert^TM^ medium overlaid with SAGE^TM^ Oil for Tissue Culture (CooperSurgical, Trumbull, CT, USA), usually 5 COC’s per well.

### Oocyte fertilization and embryo culture

Semen samples were collected or thawed on the day of oocyte retrieval. Samples were evaluated according to World Health Organization standardized guidelines (World Health Organization, 2021) and prepared by density gradient centrifugation (ORIGIO gradient™, CooperSurgical, Tumbull, CT, USA) or ZyMōt^TM^ Multi 850μL Sperm Separation Device (CooperSurgical^®^, Trumbull, CT, USA).

Insemination was performed with either conventional insemination (IVF) or intracytoplasmic sperm injection (ICSI) when indicated. IVF was performed 4-6 hours after oocyte retrieval. The procedure involved using an insemination stock of 100 x 10^3^ motile sperm/well and adding cumulus-oocyte-complexes (COCs). For ICSI, oocytes were denuded 2 hours post retrieval and sperm injection was performed approximately 2 hours later. Cumulus cells were removed using 80 IU/mL hyaluronidase (ORIGIO^®^ SynVitro^TM^ Hyadase, CooperSurgical^®^, Trumbull, CT, USA). Denuded oocytes were assessed for maturity and those scored as metaphase II (MII) underwent ICSI.

Fertilization was assessed 16-18 hours post insemination. 2PN zygotes were cultured in SAGE^TM^ 1-Step^TM^ culture medium (CooperSurgical^®,^ Trumbull, CT, USA) overlaid with SAGE^TM^ Oil for Tissue Culture (CooperSurgical, Trumbull, CT, USA). Incubators were set to 37°C, 6% CO_2_, and 5% O_2_. Embryos were assessed for developmental stage and morphological quality on days 5 and 6 post-insemination. Blastocysts were assigned a Grade of 1, 2, or 3 based on expansion, appearance, and number of cells. Grade 1 embryos were considered top quality and Grade 2 embryos were classified as good quality. Equivalence of grades to Gardner scores is presented in **Supplementary Table 1.**

### Embryo vitrification and warming

#### Traditional embryo vitrification

Blastocysts were collapsed either manually by aspiration using 175 or 275µm The Stripper^®^ tips (CooperSurgical, Trumbull, CT, USA) or using laser (RI Saturn 5^TM^ Active Laser System, CooperSurgical, Trumbull, CT, USA) and vitrified on D5 or D6 of development on VitriFit^TM^ cryo-devices (CooperSurgical, Trumbull, CT, USA). Two separate vitrification protocols were employed as part of the study. Until May 2023, blastocysts were vitrified using the Sydney IVF^TM^ Blastocyst Vitrification Kit (Cook Medical Australia, Brisbane, QLD, Australia) according to the instructions for use. Blastocysts were placed in solution 1 (10mM HEPES solution) for up to 10 minutes, then moved to solution 2 (8% ethylene glycol, 8% DMSO) for 2 minutes. Blastocysts were then exposed to solution 3 (16% ethylene glycol, 16% DMSO and 0.68M trehalose) for 20-30 seconds, before being loaded onto the VitriFit^TM^ cryo-device (CooperSurgical, Trumbull, CT, USA) and plunged into liquid nitrogen.

#### Blastocyst vitrification - shortened protocol

From May 2023, embryos were vitrified using SAGE^TM^ vitrification media kit (CooperSurgical, Trumbull, CT, USA). Solutions were brought to room temperature (20-25°C) prior to use. Blastocysts were collapsed and transferred to the ES (7.5% ethylene glycol, 7.5%% DMSO) for 2 minutes. Blastocysts were then transferred to VS (15% ethylene glycol, 15% DMSO) for 20-30 seconds, loaded onto the VitriFit^TM^ cryo-device (CooperSurgical, Trumbull, CT, USA) and plunged into liquid nitrogen.

#### Traditional multi-step warming

Multi-step warming was used prior to July 2024. Dishes were prepared for warming on the morning of the day of transfer. Solution 1 from the warming kit (SAGE^TM^ Vitrification Warming Kit, CooperSurgical, Trumbull, CT, USA) was warmed at 37 degrees for 30 mins whilst solution 2 and MOPS were brought at room temperature (20-25°C). Immediately prior to warming, 0.5mL of each solution was pipetted into an individual well of a 4-well dish (Nunc^®^,Thermo Fisher Scientific, Scoresby, VIC, Australia).

The cap was removed from the cryodevice and the curved loading area of the carrier quickly moved to the well containing WS1 (1.0M sucrose solution) on a heated microscope stage (37°C). Subsequent steps were performed at room temperature. After 1 minute the blastocyst was moved to the well containing WS2 (0.5M sucrose). After a further 3 minutes, the blastocyst was moved through 2 wells of MOPS solution (MOPS buffered solution of modified HTF), 5 minutes in each. The blastocyst was then transferred to the culture dish and washed through multiple drops of culture medium before being placed in a 20µL culture drop (SAGE^TM^ 1-Step^TM^, CooperSurgical, Trumbull, CT, USA). Culture was performed in incubators running at 37°C, 5% CO_2_, 6% O_2_, in dishes equilibrated overnight with a 1.0mL oil overlay (SAGE^TM^ Oil for Tissue Culture, CooperSurgical, Trumbull, CT, USA). Transfer was performed after 1-4hrs of post-warming culture.

#### One-step warming

One-step warming was used from July 2024 onwards. Dishes were prepared the morning of the day of transfer immediately prior to warming. WS1 1M sucrose solution (SAGE^TM^ Vitrification Warming Kit, CooperSurgical, Trumbull, CT, USA) was incubated at 37°C in a non-gassed environment for a minimum of 30 minutes. A 4-well dish (Nunc^®^, Thermo Fisher Scientific^®^, Scoresby, AUS) was prepared, with a single well of 0.5mL of warmed WS 1M sucrose solution.

The cap was removed from the cryodevice and the curved loading area of the carrier quickly moved to the well containing the warmed WS 1M sucrose solution. After 1 minute the blastocyst was transferred to the culture dish, as previously described Culture conditions were as previously described and transfer was performed after 1-4hrs.

### Endometrial preparation and FET

Frozen cycle programming and FET were performed in line with the clinic groups’ standard protocols and accepted clinical practice. There were no changes in clinic practices for programming and management of frozen cycles during the study period. Additionally, there were no changes in staff involved in performing embryo transfer or their transfer technique.

Most cases were natural cycle frozen embryo transfers. This approach was used for ovulatory patients with regular cycles, with warming and transfer occurring 6-7 days post-ovulation. In a small number of cases a “modified” natural cycle was employed, with an agonist or antagonist ovulation trigger, and warming and transfer 5-7 days later. Medicated cycles were used for patients who were anovulatory or with irregular cycles. Most used letrozole for mild stimulation and r-hCG (Ovidrel^®^, Merck Healthcare KGaA, Darmstadt, Germany) as the ovulation trigger, with warming and transfer 5-7 days post-trigger. Where clinically indicated – usually in cases where endometrial thickness may be sub-optimal - HRT protocols were used, with warming and transfer around 5 days after starting progesterone for luteal support.

ET was performed using ultrasound guidance, with the embryo placed 1-1.5cm from the uterine fundus. The Wallace^®^ Sure-Pro^®^ (CooperSurgical, Trumbull, CT, USA) or COOK^®^ Soft-Pass^TM^ with EchoTip^®^ (Cook Medical Australia, Brisbane, QLD, Australia) embryo replacement catheters were used for single-stage transfers. The Wallace^®^ SurePro^®^ Ultra Embryo Replacement Catheter with Obturator (CooperSurgical, Trumbull, CT, USA) or COOK^®^ Guardia™ Access ET Embryo Transfer Catheter (Cook Medical Australia, Brisbane, QLD, Australia) were used for elective two-stage transfers, while the Wallace^®^ Sure-Pro^®^ Ultra Embryo Replacement Catheter with Stylet (CooperSurgical^®^, Trumbull, CT, USA) was used for difficult two-stage transfers.

### Statistical analysis

Cycles were categorized into three groups based on the combination of vitrification and warming techniques: traditional vitrification with multi-step warming (S/S), short vitrification with multi-step warming (F/S), and short vitrification with single-step warming (F/F). As current standard of care, group S/S was used as the reference.

Survival post-warming was defined as preserved integrity of at least 70% of the blastomeres following 1-2 hours of recovery in culture. Embryos were grouped by grade at freezing as G1 -top- or G2 –good-, using the classification reported in **Supplementary Table 1**. Biochemical pregnancy was defined as the detection of 50 IU/L bHCG in blood 10 days post transfer. Clinical pregnancy was defined as the presence of a foetal heartbeat or foetal sac. Obstetric and perinatal data collected included pregnancy outcome (including date of live birth or date pregnancy ended), if caesarean section was performed, sex and weight of the baby, and any complications for the mother or baby.

Statistical analyses were performed in R (version 4.4.3) to examine the relationship between vitrification and warming protocols and clinical outcomes. To model the relationship between the combinations of vitrification and warming protocols and pregnancy outcomes while accounting for the non-linear effect of age, a Generalized Additive Models (GAM) was used. The model controlled for additional confounders such as embryo quality and number of transferred blastocysts. To account for the non-independence of multiple embryos from the same patient, a random intercept was included for patient ID. Model estimation was performed using restricted maximum likelihood (REML). A level of 0.05 was selected to determine statistical significance in all cases. Two-tailed chi-square with Yates correction was used to examine obstetric and perinatal outcomes; p<0.05 was considered significant.

## RESULTS

### Shortened vitrification and overall reproductive results

The combination of vitrification and warming protocols provided three study groups for outcomes from embryo survival to live birth: traditional vitrification with multi-step warming (S/S), short vitrification with multi-step warming (F/S), and short vitrification with single-step warming (F/F). This analysis included a total of 3603 blastocysts warmed, and 3335 blastocysts transferred in 3168 procedures for 2170 unique patients. The overall LBR was 33.1% (1050/3168) with female age 35.2±4.5 (range: 22-46).

A total of 751 blastocysts were transferred to 555 patients in group S/S, 991 were transferred to 734 patients in the group F/S, and 1593 were transferred to 1126 patients in group F/F. Blastocyst survival rates were 99.1%, 99.0% and 99.3% for groups S/S, F/S and F/F respectively. The biochemical pregnancy rates were 46.2% for S/S, 49.6% for F/S, and 43.9% for F/F. Clinical pregnancy rates were 43.1% for group S/S, 46.1% for group F/S, and 40.8% for group F/F. Live birth rates were 33.6% for group S/S, 36.8% for group F/S, and 30.6% for group F/F.

When controlling for maternal age, embryo quality and the number of blastocysts transferred, survival rate and biochemical, clinical pregnancy and live birth rates were comparable across groups (all p>0.54). As expected, maternal age at vitrification showed a significant non-linear correlation with live birth rates (p<0.001 for all groups).

Clinical outcome data were collected on 1367/1471 (92.9%) of transfer cycles with an initial positive βHCG, including 1358 confirmed intrauterine pregnancies and 1050 livebirths. The overall twin pregnancy rate was 3.8% (n=50) – 2.4% in single embryo transfers (31/1269) and 23.0% (20/87) in double embryo transfers. A single triplet pregnancy was recorded from a single embryo transfer in the group F/F.

Analysis of 1279 single embryo transfers with a positive βHCG, and known outcome data, showed a 10.3% (n=132) pregnancy loss rate prior to 7 weeks and an 8.3% (n=95) clinical loss rate – these outcomes were comparable across groups (all p>0.05). Detailed description of outcomes is presented in **Table 2**.

**Table 2:**
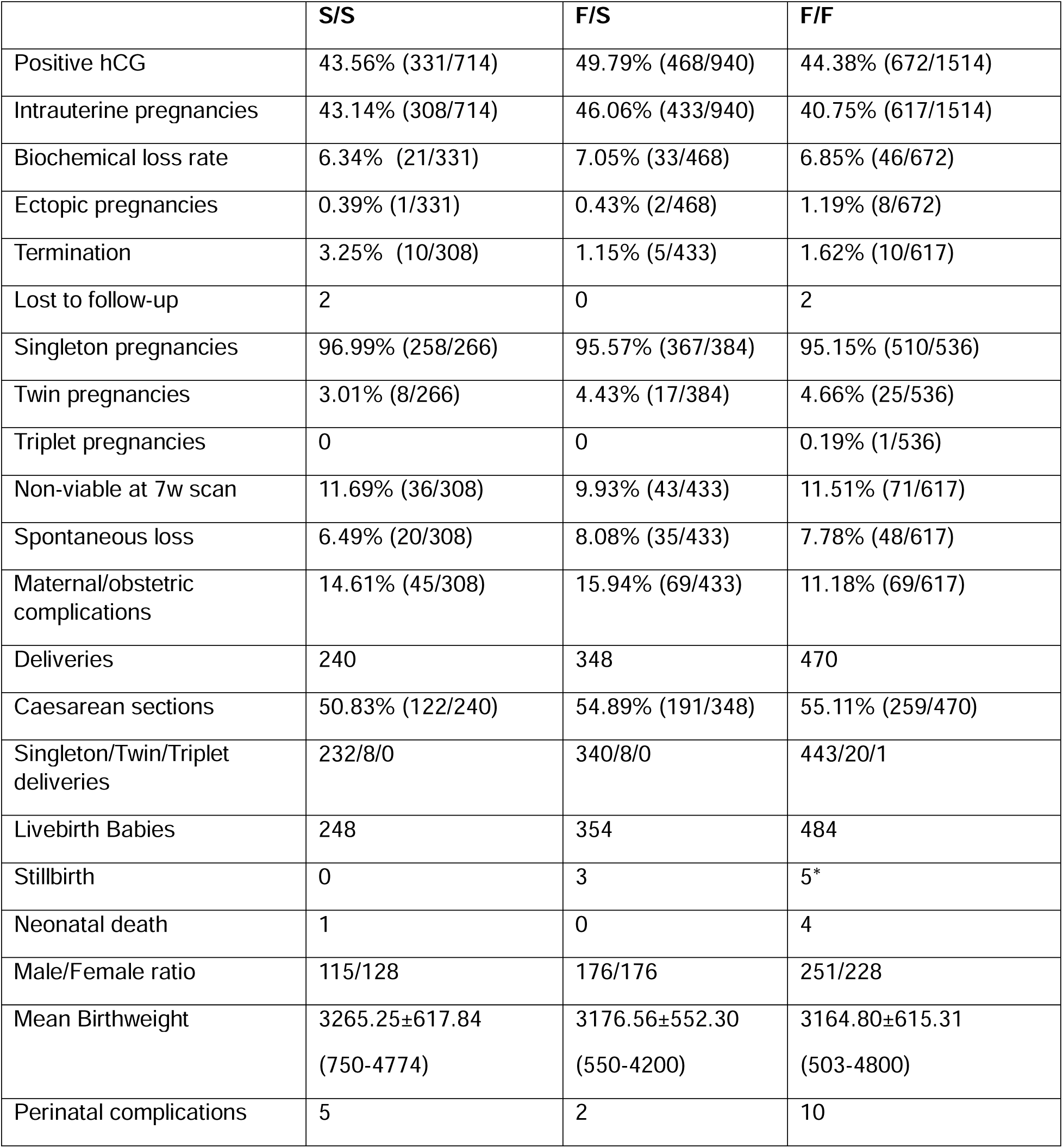
Obstetric and perinatal outcomes in the reference group (S/S = traditional vitrification/multi-step warming) and control groups (F/S = short vitrification/multi-step warming, F/F = short vitrification/one-step warming). *Includes the 3 babies from a triplet pregnancy.

|  | S/S | F/S | F/F |
| --- | --- | --- | --- |
| Positive hCG | 43.56% (331/714) | 49.79% (468/940) | 44.38% (672/1514) |
| Intrauterine pregnancies | 43.14% (308/714) | 46.06% (433/940) | 40.75% (617/1514) |
| Biochemical loss rate | 6.34% (21/331) | 7.05% (33/468) | 6.85% (46/672) |
| Ectopic pregnancies | 0.39% (1/331) | 0.43% (2/468) | 1.19% (8/672) |
| Termination | 3.25% (10/308) | 1.15% (5/433) | 1.62% (10/617) |
| Lost to follow-up | 2 | 0 | 2 |
| Singleton pregnancies | 96.99% (258/266) | 95.57% (367/384) | 95.15% (510/536) |
| Twin pregnancies | 3.01% (8/266) | 4.43% (17/384) | 4.66% (25/536) |
| Triplet pregnancies | 0 | 0 | 0.19% (1/536) |
| Non-viable at 7w scan | 11.69% (36/308) | 9.93% (43/433) | 11.51% (71/617) |
| Spontaneous loss | 6.49% (20/308) | 8.08% (35/433) | 7.78% (48/617) |
| Maternal/obstetric complications | 14.61% (45/308) | 15.94% (69/433) | 11.18% (69/617) |
| Deliveries | 240 | 348 | 470 |
| Caesarean sections | 50.83% (122/240) | 54.89% (191/348) | 55.11% (259/470) |
| Singleton/Twin/Triplet deliveries | 232/8/0 | 340/8/0 | 443/20/1 |
| Livebirth Babies | 248 | 354 | 484 |
| Stillbirth | 0 | 3 | 5* |
| Neonatal death | 1 | 0 | 4 |
| Male/Female ratio | 115/128 | 176/176 | 251/228 |
| Mean Birthweight | 3265.25±617.84<br>(750-4774) | 3176.56±552.30<br>(550-4200) | 3164.80±615.31<br>(503-4800) |
| Perinatal complications | 5 | 2 | 10 |

Obstetric and perinatal outcomes were assessed for singleton intrauterine clinical pregnancies (n=1130). Once adjusted for gestation length and sex, birthweights were comparable across groups, as was the female:male sex ratio (unadjusted p>0.05). Maternal complications were reported in 15.3% (n=183) of these pregnancies, comparable across groups. Neonatal issues ranged from common observations like jaundice, hypospadias and tongue-tie to more serious conditions, and were reported in 1.7% (n=19) of singleton pregnancies, comparable across groups (unadjusted p>0.05). Obstetric and perinatal outcome data are outlined in **Supplementary Table 2**.

### Advanced maternal age and vitrification/warming outcomes

In women ≥35yrs, F/S live birth rates seemed to decline less than S/S and F/F. Given this observation, a sub-analysis for women ≥35 years old at the time of vitrification (range: 35-46, n=1715 transfers; S/S =347, F/S=502, F/F=866) was performed.

Survival rates overall were 99.2% for group S/S, 99.0% for group F/S and 99.3% for group F/F. Biochemical pregnancy rates were 39.8% for group S/S, 45.6% for group F/S and 35.8% for group F/F. The clinical pregnancy rates were 37.2% for group S/S, 42.0% for group F/S and 33.0% for group F/F. Live birth rates for patients >35yrs were 26.2% in group S/S, 31.1% in group F/S, and 21.6% in group F/F.

No meaningful non-linearity or age association was observed for survival rates across combinations of vitrification and warming protocols (all p>0.76). The reproductive outcomes showed the expected decline in outcome probability with advancing age for all groups. While S/S and F/F were comparable across outcomes in the patients ≥35yrs after adjustment (all p>0.05), F/S showed higher adjusted odds of biochemical pregnancy (OR 1.35, [1.01-1.81], p=0.042), clinical pregnancy (OR 1.32, [0.98-1.77], p=0.069) and live birth (OR 1.42, [1.02-1.98], p=0.038) compared to S/S.

Predicted probabilities from the main GAMs at selected ages are presented in **Supplementary Table 3.**

### Embryo quality and vitrification/warming outcomes

Blastocysts were divided into two groups - top quality or good quality - based on morphological grade. Blastocysts classified as G1 were considered top quality (S/S n=223, F/S n=337, F/F n=457), while those classified as G2 were considered good quality (S/S n=528, F/S n=654, F/F n=1136; **Supplementary Table 1**). G1 and G2 blastocysts were compared for survival, biochemical, clinical, and livebirth rates across all protocol combinations.

Survival rates for G1 blastocysts were 98.3% in group S/S, 98.1% in group F/S, and 98.8% in group F/F. The survival rate for G2 blastocysts was 99.4% in group S/S, 99.4% in group F/S, and 99.5% in group F/F.

Biochemical pregnancy rates for G1 blastocysts were 59.0% in group S/S, 53.8% in group F/S, and 56.4% in group F/F, while for G2 they were 40.6%, 47.4% and 38.9% respectively. Clinical pregnancy rates for G1 blastocysts were 54.4% in group S/S, 51.6% in group F/S, and 52.7% in group F/F, while for G2 they were 38.2%, 43.2% and 36.0% respectively. Finally, live birth rates for G1 blastocysts were 47.5% in group S/S, 41.2% in group F/S, 42.5% in group F/F, while for G2 were 27.6%, 34.5%, and 25.9% respectively.

No differences were observed in survival between the two quality groups (p=0.136). As expected, a significantly lower probability of pregnancy and live birth was observed for G2 blastocysts compared to G1 across all test groups (47% lower odds of live birth, p<0.001). However, within the G2 group, blastocyst undergoing fast vitrification with multi-step warming showed significantly higher biochemical pregnancy rates (OR 1.62 [1.05–2.51], p=0.030) and live birth rates (OR 1.78 [1.12–2.83], p=0.015) compared to G2 blastocysts in the S/S group. No equivalent interaction was observed for F/F (biochemical pregnancy: p=0.721; live birth: p=0.423).

### Day of vitrification and vitrification/warming outcomes

Embryos were either vitrified on Day 5 or Day 6 based on their development to blastocyst. Transfers of blastocysts vitrified on Day 5 (Group S/S n=623, Group F/S n=864, Group F/F n=1362) and Day 6 (Group S/S n=91, Group F/S n=76, Group F/F n=152) of development were compared for survival, biochemical, clinical, and livebirth rates across all protocol combinations. Survival rate for D5 blastocysts was 99.1% for group S/S, 99.0% for group F/S and 99.3% for group F/F. D6 survival rate was 98.9% for group S/S, 98.8% for group F/S and 99.5% for group F/F.

Biochemical pregnancy rates for D5 blastocysts were 48.3% for group S/S, 50.6% for group F/S and 46.1% for group F/F, while for D6 were 31.9%, 38.2% and 23.7%, respectively. The clinical pregnancy rates for D5 blastocysts were 45.3% for group S/S, 47.0% for group F/S and 43.0% for group F/F, while for D6 were 28.6%, 35.5% and 21.1%, respectively. Live birth rates for D5 blastocysts were 35.8% in group S/S, 38.3% in group F/S, and 32.7% in group F/F, while for D6 embryos were 18.7%, 19.7% and 15.8% respectively.

Overall, D6 blastocysts had significantly worse reproductive outcomes than D5 when controlling for age and number of transferred blastocysts (51% lower chances of live birth, p=0.018). This impact was consistent across protocols combinations, with no differences in biochemical (F/S p=0.477, F/F p=0.210) and clinical pregnancy (F/S p=0.366; F/F p=0.240) and live birth rate (F/S p=0.916; F/F p=0.722).

### Male factor infertility and vitrification/warming outcomes

In this study cohort, ICSI was only performed in cases of a male factor infertility diagnosis. This strict network policy allowed comparison of ICSI-derived blastocyst transfers (S/S n=271, F/S n=403, F/F n=647) to IVF (S/S n=443, F/S n=537, F/F n=867) as a proxy measure for male infertility. IVF and ICSI derived blastocyst groups were for survival, biochemical, clinical, and livebirth rates across all protocol combinations.

Survival rates in the ICSI group were 99.0% for group S/S, 99.3% for group F/S and 99.3% for group F/F and were 99.2%, 98.7%, and 99.3% respectively in IVF cycles. Biochemical pregnancy rates were 43.9% for group S/S, 48.9% for group F/S and 42.3% for group F/F in ICSI cycles and 47.6%, 50.1%, and 45.0% respectively in IVF cycles. Clinical pregnancy rates were 42.1% for group S/S, 46.4% for group F/S and 38.9% for group F/F in ICSI cycles and 43.8%, 45.8%, and 42.1% respectively in IVF cycles. Live birth rates in ICSI cycles were 34.3% for group S/S, 36.7% for group F/S and 28.9% for group F/F, and in IVF cycles were 33.2%, 36.9%, and 31.9% respectively.

Overall, rates for survival (p=0.834), biochemical (p=0.428) and clinical pregnancy (p=0.813), and live birth (p=0.567) were comparable between ICSI and IVF derived embryos across vitrification and warming protocols when controlling for key variables (all adjusted p>0.22 for survival, biochemical and clinical pregnancy, and live birth rate across study groups).

## DISCUSSION

We show here, in the largest and most comprehensive cohort reported so far, that blastocyst vitrification can be significantly shortened to 2.5 minutes in total with no prejudice to viability and competence of the embryos to give rise to sustained pregnancies, including obstetrical and perinatal outcomes.

Effective vitrification hinges on the delivering and subsequently removing of cryoprotective agents (CPAs) while minimizing exposure to high CPAs concentrations, osmotic stress, and toxicity. CPA toxicity may produce subtle, embryo-specific effects across multiple cellular pathways (Best, 2015; Verheijen *et al*., 2019; Schiewe *et al*., 2024; Zhang *et al*., 2024); to mitigate these risks, two main strategies have been devised: first, stepwise equilibration, employed in both traditional and shortened protocols, reduces osmotic stress (Karlsson *et al*., 2014); second, shortened CPAs exposure to the minimum required for vitrification limits their toxicity on cellular components such as membranes and mitochondria (Jones *et al*., 2004; Huebinger, 2018) —this being a defining feature of shortened protocols. In conventional blastocyst vitrification, cells are equilibrated in CPAs for up to 15 minutes. However, earlier studies demonstrated that exposure times as short as 1 minute can lead to sufficient

dehydration and CPA penetration for survival and development (Gallardo *et al*., 2019). This shortened approach has been validated in oocytes pre-clinically (Schiewe *et al*., 2024; Wozniak *et al*., 2024) and clinically (Aydin *et al*., 2024; Gudkova *et al*., 2024), and potentially extended to blastocysts (Gallardo *et al*., 2019; Schiewe *et al*., 2024), with comparable outcomes to traditional protocols (Kaskar *et al*., 2023; Arkfeld *et al*., 2025), consistent with our findings. Further, in a bovine model of blastocyst vitrification, shorter exposure to ES correlated with increased survival and hatching rates post-warming, and higher total and trophectoderm cell number in D7 blastocysts reaching the hatching stage within 24hrs; additionally, the shorter equilibration resulted in a significantly lower apoptotic cell count (Martínez-Rodero *et al*., 2021).

We further identified an intriguing and positive effect of short vitrification combined with multi-step warming on reproductive outcomes of AMA patients up and including live birth rates. This observation was unexpected and needs further validation before being considered in clinical practice. A possible explanation may reside in a higher susceptibility of blastocysts from advanced maternal age patients to CPA toxicity and osmotic stress. As such, shortened vitrification protocols limit the exposure to CPAs, reducing the risk of permeability changes and/or damage to cell membranes. Pairing this with multi-step warming may help ensure that all CPAs are removed during the warming process, as excess CPA loading during vitrification may be unable to effectively leave the cells during warming. This aligns with the observed over-hydration of blastocyst cells, overexpansion of the blastocoele, membrane rupture and cell necrosis with single step warming compared to traditional protocols (Ezoe *et al*., 2025).

We recognize some limitations to this study: most importantly, the retrospective nature of the data does not allow for adequate controlling of unaccounted for variables. Further, the inclusion of data from clinics operating with comparable standardized protocols allowed to control for variation in protocols and practices but may limit the generalizability of our results.

In conclusion, the non-inferiority of shortened vitrification and warming protocols to traditional protocols in terms of clinical, obstetric and perinatal outcomes brings about operational efficiency, a stronger chain of custody, and lower risk of error as the number of frozen embryo cycles continue to rise globally. Further, we provide initial evidence that the combination of shortened vitrification with traditional multi-step warming may improve outcomes in patients of advanced maternal age.

## DECLARATION

### Data availability

The data underlying this article are available upon reasonable request to the corresponding author.

## Acknowledgements

We would like to thank Filippo Zambelli of TRT Consulting for statistical support.

## CRediT Authorship Contribution Statement

**Emma R. Ebinger:** Conceptualization, Data validation, Supervision, Writing. **Helena Misurac:** Data curation. **Colleen Lynch:** Writing review & editing, data visualization. **Vrunda Desai**: Review, Resources. **Nadia De Rosa:** Writing. **Rita Vassena:** Writing review & editing, Methodology, Conceptualization. **Paul J. Atkinson:** Writing.

## Funding

None

## Conflict of interest

Authors CL, VBD and RV are employees of CooperSurgical Inc..

## Notes

### Competing Interest Statement

The authors have declared no competing interest.

### Author Declarations

Ethics committee/IRB of ADORA Fertility gave ethical approval for this work

## REFERENCES

Arkfeld CK, Vagios S, Jiang VS, Minis E, Bormann CL. COMPARABLE LIVE BIRTH AND CLINICAL OUTCOMES FROM BLASTOCYSTS VITRIFIED WITH A SHORTENED 3-MINUTE PROTOCOL VERSUS A STANDARD METHOD. Fertil Steril 2025;124:E5.

Aydin B, Hudkova D, Maggiotto G, Kal NS, Osmanllari U, Unsal E, Baltaci V, Aktuna S, Yonar D, Dorofeyeva U, et al. Human oocyte survival, early embryo development, metabolic fingerprinting, and pregnancy outcomes following ultra-rapid or standard vitrification and thawing. Human Reproduction 2024;39:deae108.351.

Best BP. Cryoprotectant Toxicity: Facts, Issues, and Questions. Rejuvenation Res 2015;18:422–436.

Dyer S, Chambers GM, Jwa SC, Baker VL, Banker M, Mouzon J de, Elgindy E, Fu B, Ishihara O, Kupka MS, et al. International Committee for Monitoring Assisted Reproductive Technologies world report: assisted reproductive technology, 2019. Fertil Steril 2025;124:679–693.

Ebinger ER, Misurac H, Giovannini AM, Desai VB, Rosa N De, Vassena R, Atkinson PJ. Efficacy and safety of a one-step warming protocol of vitrified blastocyst stage embryos. F S Rep [Internet] 2026;Available from: https://linkinghub.elsevier.com/retrieve/pii/S2666334126000620.

Ezoe K, Miki T, Fujiwara N, Kato K. Influence of the shortened warming protocol on human blastocyst viability: an in-vitro experimental study. Reprod Biomed Online 2025;50:104454. Elsevier Ltd.

Fahy GM, Rall WF. Vitrification: an overview. 2007;

Gallardo M, Saenz J, Risco R. Human oocytes and zygotes are ready for ultra-fast vitrification after 2 minutes of exposure to standard CPA solutions. Sci Rep 2019;9:15986. Nature Publishing Group.

Gudkova D, Aydin B, Dorofeyeva U, Yetim I, Kal NS. CLINICAL AND METABOLIC EFFECTS OF ULTRA RAPID VITRIFICATION AND THAWING ON OOCYTE AND EMBRYO VIABILITY. Fertil Steril 2024;122:.

Huebinger J. Modification of cellular membranes conveys cryoprotection to cells during rapid, nonequilibrium cryopreservation. PLoS One 2018;13:e0205520.

Jones A, Blerkom J Van, Davis P, Toledo AA. Cryopreservation of metaphase II human oocytes effects mitochondrial membrane potential: Implications for developmental competence. Human Reproduction 2004;19:1861–1866.

Karagianni M, Papatheodorou A, Oraiopoulou C, Papadopoulou MI, Thivaiou L, Katsakoglou N, Christoforidis N, Chatziparasidou A. Effect of a one-step fast warming protocol on reproductive outcomes of vitrified-warmed blastocysts. Fertil Steril 2025;124:1272–1282. Elsevier Inc.

Karlsson JOM, Szurek EA, Higgins AZ, Lee SR, Eroglu A. Optimization of cryoprotectant loading into murine and human oocytes. Cryobiology 2014;68:18–28.

Kaskar K, Sieren KR, Brabner VM, Huneke BR, Vermilyea L, VerMilyea M, Silverberg K. EFFECT OF REDUCING THE VITRIFICATION AND WARMING TIMES ON BLASTOCYST SURVIVAL AND PREGNANCY RATES. Fertil Steril 2023;120:E143.

Liebermann J, Hrvojevic K, Hirshfeld-Cytron J, Brohammer R, Wagner Y, Susralski A, Jasulaitis S, Chan S, Takhsh E, Uhler M. Fast and furious: pregnancy outcome with one-step rehydration in the warming protocol for human blastocysts. Reprod Biomed Online 2024;48:103731.

Manns J, Patrick JL, Katz I, Holt T, Katz SL, Taylor TH. Clinical validation of a new, ultrafast warming protocol, resulting in equivalent implantation rates and significant time savings versus routine warming protocol, a prospective randomized control. Fertil Steril 2022;118:.

Martínez-Rodero I, García-Martínez T, Ordóñez-León EA, Vendrell-Flotats M, Hidalgo CO, Esmoris J, Mendibil X, Azcarate S, López-Béjar M, Yeste M, et al. A shorter equilibration period improves post-warming outcomes after vitrification and in straw dilution of in vitro-produced bovine embryos. Biology (Basel) 2021;10:142.

Muldrew K, McGann LE. The osmotic rupture hypothesis of intracellular freezing injury. Biophys J 1994;66:532–541.

Schiewe MC, Reichelderfer R, Wozniak K, Romana C De, Nordbak M, Baek K, Chung K. Ultra-fast vitrification and rapid elution of human oocytes: part I. germinal vesicle model validation. Reprod Biomed Online 2024;49:104691.

Seki S, Mazur P. Effect of warming rate on the survival of vitrified mouse oocytes and on the recrystallization of intracellular ice. Biol Reprod 2008;79:727–737.

Verheijen M, Lienhard M, Schrooders Y, Clayton O, Nudischer R, Boerno S, Timmermann B, Selevsek N, Schlapbach R, Gmuender H, et al. DMSO induces drastic changes in human cellular processes and epigenetic landscape in vitro. Sci Rep 2019;9:4641.

World Health Organization. World Health Organization. WHO laboratory manual for the examination and processing of human semen. 6th ed. WHO Press 2021;

Wozniak K, Reichelderfer R, Ghaemi S, Hupp D, Fuzesi P, Ringler G, Marrs RP, Schiewe MC. Ultra-fast vitrification and rapid elution of human oocytes: Part II – verification of blastocyst development from mature oocytes. Reprod Biomed Online 2024;49:104690.

Zhang K, Lan T, Lin F, Liu R, He Q, Gao F, Wu S, Kang J, Li H, Quan F. Effect of vitrification on protein O-GlcNAcylation in mouse metaphase II oocytes. Reproduction 2024;167:e230143.

