## Supplemental Table for "Shortened blastocyst vitrification achieves live birth rates comparable to standard protocols: an analysis of 3168 cryotransfers"

**SUPPLEMENTARY TABLES**

| **Grade** | **Corresponding Gardner Grade** |
| --- | --- |
| G1 | 3AA, 4AA, 5AA, 6AA |
| G2 | 3AB, 3BA, 4AB, 4BA, 5AB, 5BA, 6AB, 6BA, 3BB, 4BB, 5BB, 6BB |

**Supplementary Table S1**: Conversion of embryo morphological grades used in the reported cases (G1 and G2) and Gardner’s grades.

| Type | Description | S/S | F/S | F/F | TOTAL |
| --- | --- | --- | --- | --- | --- |
| Maternal/ Obstetric | Gestational diabetes | 24 | 38 | 22 | 84 |
|  | Placenta Praevia | 2 | 2 | 6 | 10 |
|  | Post-Partum Haemorrhage | 2 | 1 |  | 3 |
|  | Placental insufficiency/IUGR | 2 | 3 | 1 | 6 |
|  | Hypertension | 2 | 4 | 9 | 15 |
|  | Pre-eclampsia | 8 | 11 | 18 | 37 |
|  | Hyperemesis | 3 | 6 | 6 | 15 |
|  | Cholestasis | 3 | 2 | 4 | 9 |
|  | Threatened Pre-term Labour | 3 | 1 | 1 | 5 |
|  | Preterm Prelabour Rupture of Membranes | 2 |  |  | 2 |
|  | Placenta Accreta | 1 | 1 | 1 | 3 |
|  | Anaemia | 2 | 4 | 1 | 7 |
|  | Polyhydramnios | 1 | 1 |  | 2 |
|  | Prolonged bleeding in pregnancy |  | 2 |  | 2 |
|  | Antepartum Haemorrhage |  | 1 | 2 | 3 |
|  | Placental Abruption |  | 1 | 3 | 4 |
|  | Pre-labour Rupture of Membranes |  |  | 1 | 1 |
| Perinatal | Jaundice | 1 |  | 1 | 2 |
|  | NICU for O_2_ | 1 |  |  | 1 |
|  | ICU/SCBU for 3wks due to preterm birth (35+4wks) | 1 |  |  | 1 |
|  | Hypoglycaemia | 1 |  | 2 | 3 |
|  | Asyncliticism | 1 |  |  | 1 |
|  | Tongue Tie |  | 1 |  | 1 |
|  | Hip Dysplasia |  | 1 |  | 1 |
|  | Moderate pulmonary stenosis |  |  | 1 | 1 |
|  | underweight |  |  | 1 | 1 |
|  | bilateral grade 3 brain bleed |  |  | 1 | 1 |
|  | under-developed lungs |  |  | 1 | 1 |
|  | Supraventricular tachycardia |  |  | 1 | 1 |
|  | Duplex kidney |  |  | 1 | 1 |
|  | Medium Ventricular Septal Defect |  |  | 1 | 1 |
|  | Posterior urethral valve |  |  | 1 | 1 |
|  | Laryngomalacia |  |  | 1 | 1 |

**Supplementary Table S2**: Reported maternal/obstetric and perinatal complications in the reference group (S/S = traditional vitrification/multi-step warming) and control groups (F/S = short vitrification/multi-step warming, F/F = short vitrification/one-step warming).

| **Age** | **Study group** | **Biochemical model** | **Clinical model** | **Live birth model** |
| --- | --- | --- | --- | --- |
| 32 | slow_slow | 47.0% | 37.0% | 25.2% |
| 32 | fast_fast | 50.7% | 40.9% | 28.5% |
| 32 | fast_slow | 48.6% | 38.6% | 27.6% |
| 35 | slow_slow | 41.8% | 32.4% | 21.0% |
| 35 | fast_fast | 44.0% | 34.5% | 22.6% |
| 35 | fast_slow | 44.6% | 34.4% | 23.0% |
| 38 | slow_slow | 35.6% | 26.8% | 15.3% |
| 38 | fast_fast | 34.3% | 25.6% | 13.1% |
| 38 | fast_slow | 40.8% | 30.4% | 17.9% |
| 40 | slow_slow | 30.9% | 22.9% | 11.6% |
| 40 | fast_fast | 27.6% | 19.6% | 8.1% |
| 40 | fast_slow | 38.3% | 27.9% | 14.2% |
| 42 | slow_slow | 26.3% | 19.0% | 8.5% |
| 42 | fast_fast | 21.4% | 14.3% | 4.9% |
| 42 | fast_slow | 35.8% | 25.6% | 10.9% |

**Supplementary Table S3**: Predicted probabilities from the main GAMs at selected ages, holding the other model covariates at reference/modal values.
